# Profiles of bacteria isolates and their antimicrobial resistance pattern among housemaids working in communal living residences in Jimma City, Ethiopia

**DOI:** 10.1101/2023.03.23.23287624

**Authors:** Tadele Shiwito Ango, Tizita Teshome, Tesfalem Getahun, Girma Mamo, Negalgn Byadgie

## Abstract

**Background:** Bacterial pathogens continued to be the major causes of foodborne gastroenteritis in humans and remained public health important pathogens across the globe. As regards, housemaids operating inside a kitchen could be the source of infection and may transmit disease-inflicting pathogens through infected hands. Profiles of bacteria isolates and their antimicrobial resistance patterns among housemaids employed in dwellings in Ethiopia; particularly in the study area haven’t been underexplored yet.

**Objective:** A study aimed to assess the profiles of bacteria isolates and antimicrobial resistance patterns among housemaids working in communal living residences in Jimma City, Ethiopia.

**Methods:** Laboratory-based cross-sectional study design was employed among 230 housemaids from April-June 2022. Hand swabs samples from the dominant hand of the study participants were collected under sterile conditions for the segregation of commensal microbes following standard operating procedures. Then in the laboratory, the swabs were inoculated aseptically using streak-plating methods on mannitol salt agar, MacConkey agar, Salmonella-shigella agar, and Eosin Methylene Blue Agar. Then inoculated samples were incubated at 37°C for 24 hours for bacterial isolation. In addition, a set of biochemical tests was applied to examine bacterial species. Data was entered into Epidata version 3.1. All statistics were performed using SPSS^®^ statistics version 26. Descriptive analyses were summarized using frequency and percentage.

**Results:** The response rate of respondents was 97.8%. The prevalence of bacterial contaminants in the hands of housemaids who tested positive was 72% (95%CI: 66.2-77.8%). The isolated bacterial were *Staphylococcus aureus* (31.8%), *Coagulase-Negative Staphylococci* (0.9%), *Escherichia coli* (21.5%), *Salmonella* (1.3%), *Shigella* (6.7%), *Klebsiella species* (23.3%) and *Proteus species* (14.8%). The isolation rate of bacteria was significantly associated with the removal watch, ring, and bracelet during hand washing. Most isolated bacteria were sensitive to Chloramphenicol while the majority of them were resistant to Tetracycline, Gentamycin, Vancomycin, and Ceftazidime.

**Conclusions:** Hands of housemaids are important potential sources of disease-causing bacterial pathogens that would result in the potential risk of foodborne diseases. Most isolated bacteria were resistant to tested antimicrobial drugs. Everybody responsible should work practice of good hand hygiene.

## 1. Introduction

Enteric bacterial pathogens are the major causes of foodborne gastroenteritis in humans and remain public health important pathogens worldwide [**1**]. These enteric bacterial pathogens are common food-borne disease agents and persisted as a major public health worry. A study revealed that food commodities were contaminated by food handlers or housemaids [**2**]. Moreover, the majority of foodborne outbreak causative agents enter the body through the ingestion of contaminated food [**3**,**4**]. According to Banik and colleagues, foodborne illnesses occurred after those disease-causing microbes entered the food supply chain [**5**]. WHO report in 2020, showed that there were about 600 million cases and 420,000 deaths related to contaminated food in the world [**4**]. However, the problem is severe in developing countries including Ethiopia. The summary report of the Ministry of Health revealed that the annual incidences of food-borne illnesses ranged from 3.4 to 9.3% in Ethiopia [**6**].

Fecal-oral route of pathogen transmission is the major among the other methods of infection transmission for heterogeneous pathogens particularly for *Staphylococcus aureus, Coagulase-Negative Staphylococci, Klebsiella species, Proteus species, E*.*coli, Shigella, Salmonella species, V*.*Cholerae, Streptococcus pneumonia*, and others bacterial isolates [**5**,**7–14**]. Furthermore, studies revealed that bacteria were the most extensively identified infectious agent for the majority of foodborne outbreak types [**6**,**15**].

Due to the high prevalence of drug resistance pathogens, advances in infection control haven’t completely eradicated the problems [**10**]. In addition, the constant increase in AMR bacterial strains has become an important clinical problem [**16**,**17**]. They include the members of Enterobacteriaceae, and continued the increasing concern and lead to the narrowing of available therapeutic options [**17**,**18**]. Those AMRs are caused by different reasons. Thus, the microbial evolution and transmission of genetic determinants of resistance between microbes enable the spread of pathogenic bacteria [**17–21**]. In addition, the widespread and prolonged use of antibiotics leads to the emergence of resistant bacterial pathogens [**10**,**21**,**22**]. In sum, it is an emerging global challenge that results in the spread of infectious diseases that affect human populations [**21**,**23**]. Moreover, the problems of infectious diseases were worsened by the improper use of antibiotics by humans and animals which contribute to the rise of AMR globally [**10**,**16–21**,**23–25**].

Therefore, pathogenic microbes continued the challenge for healthcare systems in developing countries including Ethiopia [**24**]. Evidence from studies revealed an increasing incidence of multidrug resistance in food-borne pathogens particularly to the commonly used antimicrobial medications [**24**,**25**]. In the fact, most studies conducted were institution based such as the hospital, mass food processing, and catering establishments [**9**,**10**,**15**,**22**], still now in Ethiopia, there is no clear data or information that shows profiles of isolated bacteria and antimicrobial resistance among housemaids in dwellings, particularly in the study area. Therefore, the present study aimed to assess the profiles of bacteria isolates and their antimicrobial resistance pattern among housemaids working in communal living residences in Jimma City, Ethiopia.

## 2. Materials and Methods

### 2.1. Study area, design, and period

The present laboratory-based cross-sectional survey included 230 housemaids engaged in communal living residences in Jimma City, Jimma Zone, Oromia region, Southwest part of Ethiopia during the period from April-June, 2022. The Jimma city is located 352km southwest of Addis Ababa with geographical coordinates between 7º41’ N latitude and 36º50’ E longitudes, and it has an average altitude of 1780m above sea level. It receives a mean annual rainfall of about 1530 millimeters. The mean annual minimum and maximum temperatures of Jimma city are 14.4°C and 26.7°C respectively [**26**].

### 2.2. Data collection techniques

Data was collected by the data collectors after obtaining written informed consent using a pre-tested semi-structured questionnaire and observation designed to obtain socio-demographic data like sex, age and educational status, and other relevant data related to housemaids’ hand hygiene practices such as fingernail status, frequent handwashing, how to wash hands, use of soap and water for frequent hand washing, follow five steps to wash hands in the right way, removal of watch, ring, and bracelets during handwashing and time in second to wash hands from the study participants following their written informed consent and ethical approval of the study approved from the Institutional Review Board of the Institute of Health, Jimma University.

### 2.3. Laboratory data, analysis, and interpretation

Data on the commensal microbes from the hand swabs were collected through laboratory investigation by following the standard operating procedures (SOPs).

#### 2.3.1. Sample collection and transport

Sterile cotton swabs and 10ml saline containing sterile test tubes were prepared to collect and transport the samples. Though for the bacterial isolates from hand swabs, after handwashing participant’s dominant hand was sampled by rubbing all over the surface using sterile-moistened cotton-tipped swabs in the moistened state; and then placed/soaked in labeled 0.85% saline solution containing sterile test tubes for microbial culturing. However, notification was not delivered in advance and extra hand hygiene was not allowed during sample collection [**22**,**27**]. Swabs samples were collected by three well-trained laboratory personnel in standard aseptic procedures. Soon after collection, samples were sent to the Microbiology laboratory at the Department of Medical Microbiology at Jimma University. Then in the laboratory, the samples were enriched in nutrient broth for 24 hours to enhance the recovery of the isolates because the survival of bacteria collected can be affected by handwashing.

#### 2.3.2. Sample culture and identification

The common method to identify bacteria is through the of use selective media which can hinder or suppresses the growth of unwanted commensal microbes or the use of differential media which is easier to distinguish colonies of desired micro-organisms from other colonies growing on the same plate [**28**,**29**].

The media used in this study were prepared according to the manufacturer’s instructions. A loop full of each hand swabs sample enriched on nutrient broth was inoculated aseptically using streak-plating methods on the selective and differential (Mannitol salt agar; MacConkey agar; salmonella-shigella agar and Eosin Methylene Blue Agar) and then incubated at 37°C for 24 hours. After an incubation period, the culture plates were examined for the growth of bacteria, and the morphology of the isolates was recorded.

#### 2.3.3. Biochemical tests

The single colony of bacteria grown on selective and differential media was then subcultured into nutrient agar to determine growth patterns and for further biochemical tests. Then after obtaining pure colonies, identification of bacterial isolates was done by using standard microbiology techniques like the morphology of its colonies and a battery (set) of biochemical tests like a response on catalase, coagulase, oxidase, Simon citrate agar (SCA), urease, sulfide indole motility (SIM), Kliger Iron Agar (KIA), gas and hydrogen sulfide (H2S) generation [**14**]. The isolation and identification of bacteria from hand swabs from the hands of housemaids are shown below (**Fig 1**).

**Figure 1:**
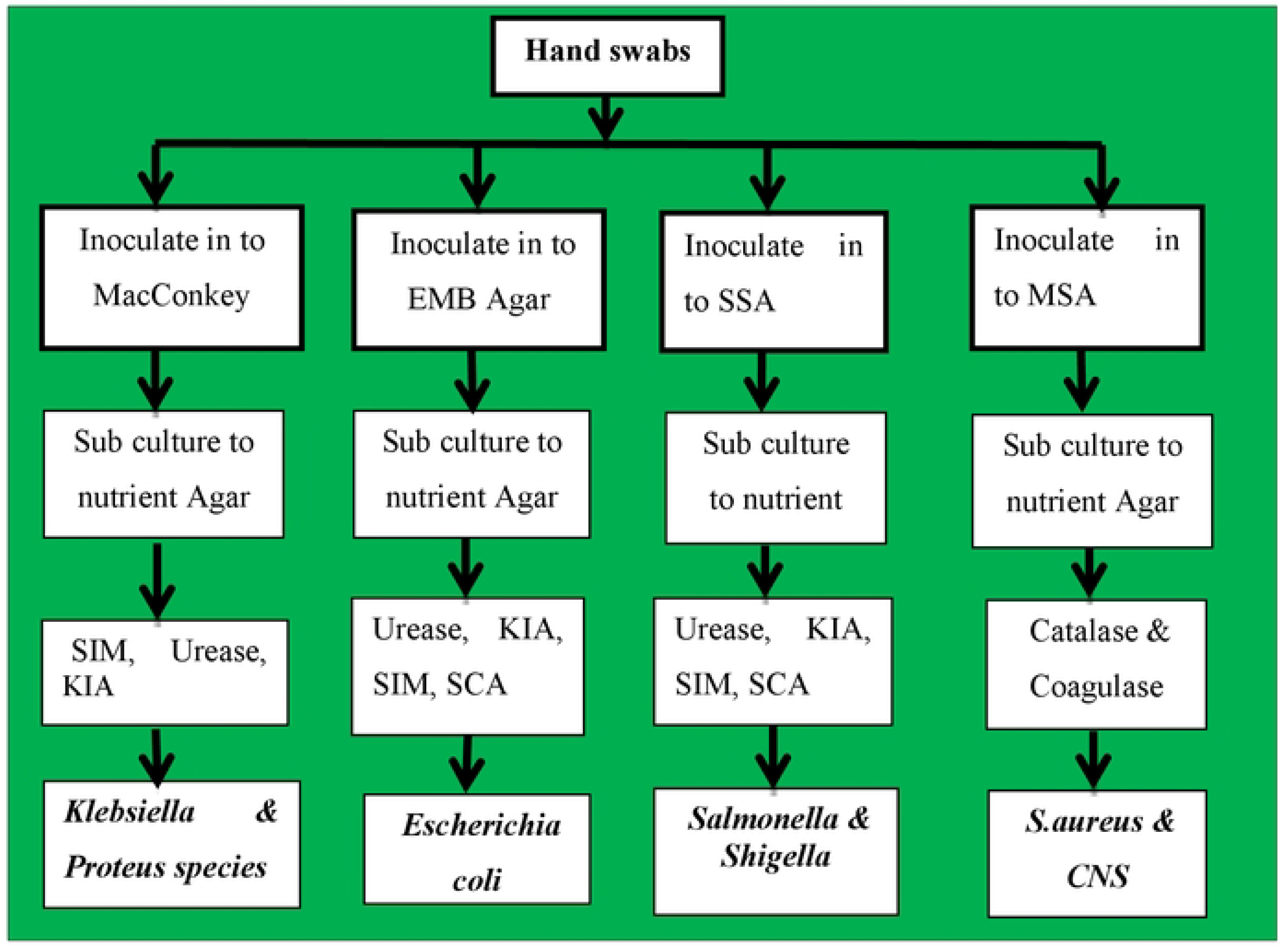
Laboratory flow chart showing bacterial isolations from hand swabs samples.

#### 2.3.4. Antimicrobial susceptibility tests

Antimicrobial susceptibility tests were performed on Muller Hinton Agar (HIMEDIA, TITAN BIOTECH LTD, Rajasthan, India) by disc diffusion method. The following antimicrobial drugs were used to test susceptibility: Tetracycline (30μg), Ceftriaxone (30μg), Chloramphenicol (30μg), Gentamicin (10μg), Vancomycin (30μg), and Ceftazidime (30μg). The selections of drugs were based on availability and pieces of literature [**14**,**30**]. The susceptibility profiles that mean sensitivity, intermediate and resistance of the bacterial isolates were interpreted according to the NCCLSs [**30**].

### 2.4. Data quality management

To control the quality of laboratory data, laboratory tests for the investigation of commensal microbes have strictly adhered to standard operating processing [**29**]. In addition, the proper functioning of the instruments utilized was checked before processing samples and the known strains of selected organisms (*S*.*aureus* ATCC12981 and *E*.*coli* ATCC25922) were used for comparison purposes amid distinguishing proof as far quality

### 2.5. Data processing and analysis

Data were edited, cleaned, and double-entered into Epidata version 3.1. All statistical calculations were performed using IBM^®^ SPSS^®^ Statistics version 26. Descriptive analyses were summarized using frequency and percentage to present in texts, tables, and figures.

### 2.6. Ethical consideration

The research was conducted after approval of ethical clearance by the IRB of Jimma University. Both oral and informed consent was sought from each respondent. The overall information obtained from study participants and their privacy was kept strictly confidential using codes. The unusual clinical finding would be linked to concerned bodies including the study participants and households. PPE was applied such as the use of a mask, the use of gloves, rubbing hands with sanitizer or alcohol, and hand washing with soap during data collection to prevent transmission of Covid-19 between the data collector to the study participants and vice-versa.

## 3. Results

### 3.1. Socio-demographic characteristics

Two hundred and twenty-five study subjects (housemaids) participated in this study. All the respondents were females. The age of study participants ranged from 18 to 36 with a mean age of 21.41±SD of (3.961). The majority, 182(81%) and 34(15%) of the study participants were between the age category of 18-30 years and age 18-24years respectively (**Fig 2**).

**Figure 2:**
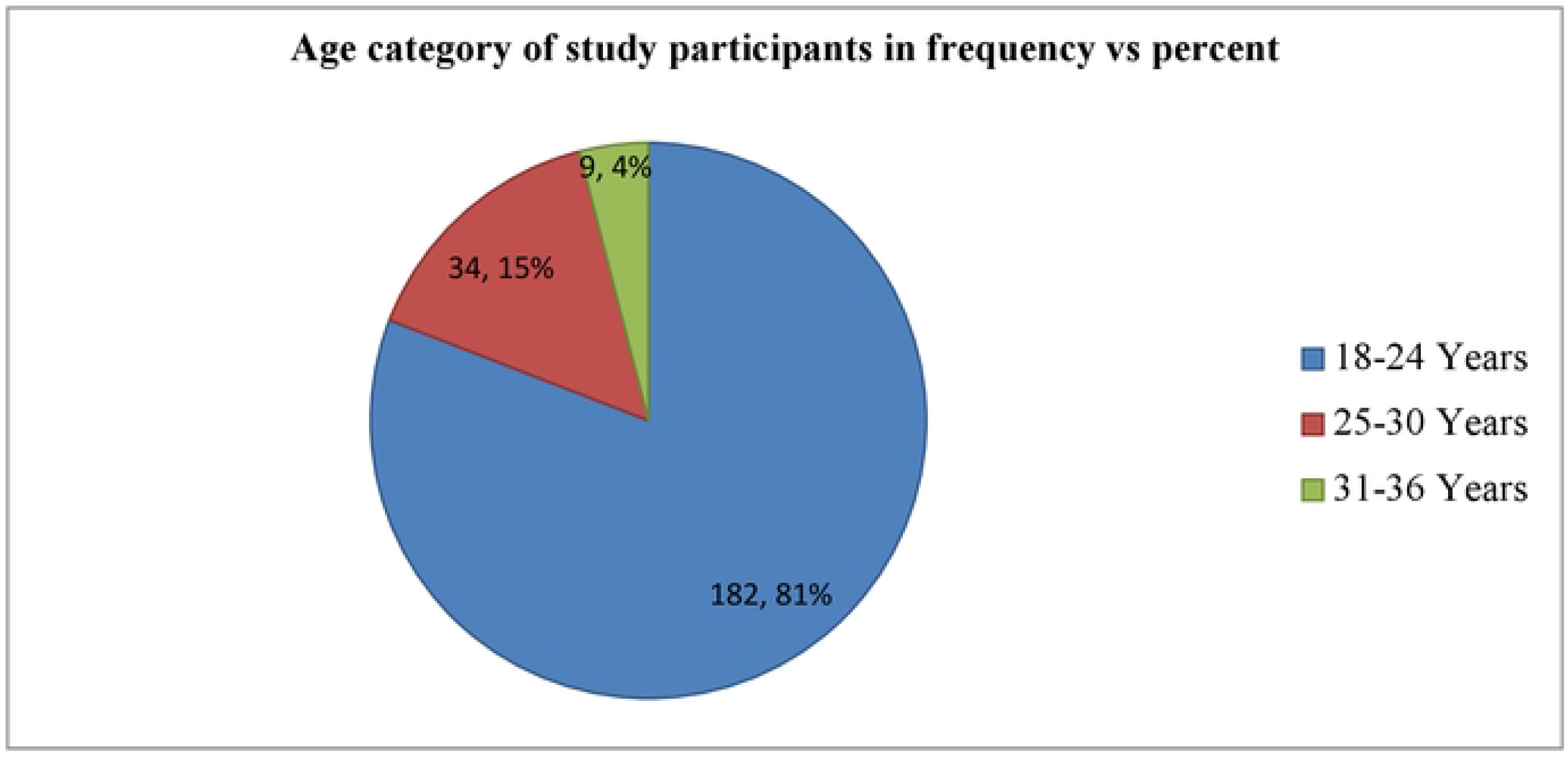
Pi chart showing the age category of housemaids (n=225) working in communal residences in Jimma City, Southwest Ethiopia, 2022.

More than half, (53%) of study participants attended primary school while 7(3%) of respondents cannot read and write (**Fig 3**).

**Figure 3:**
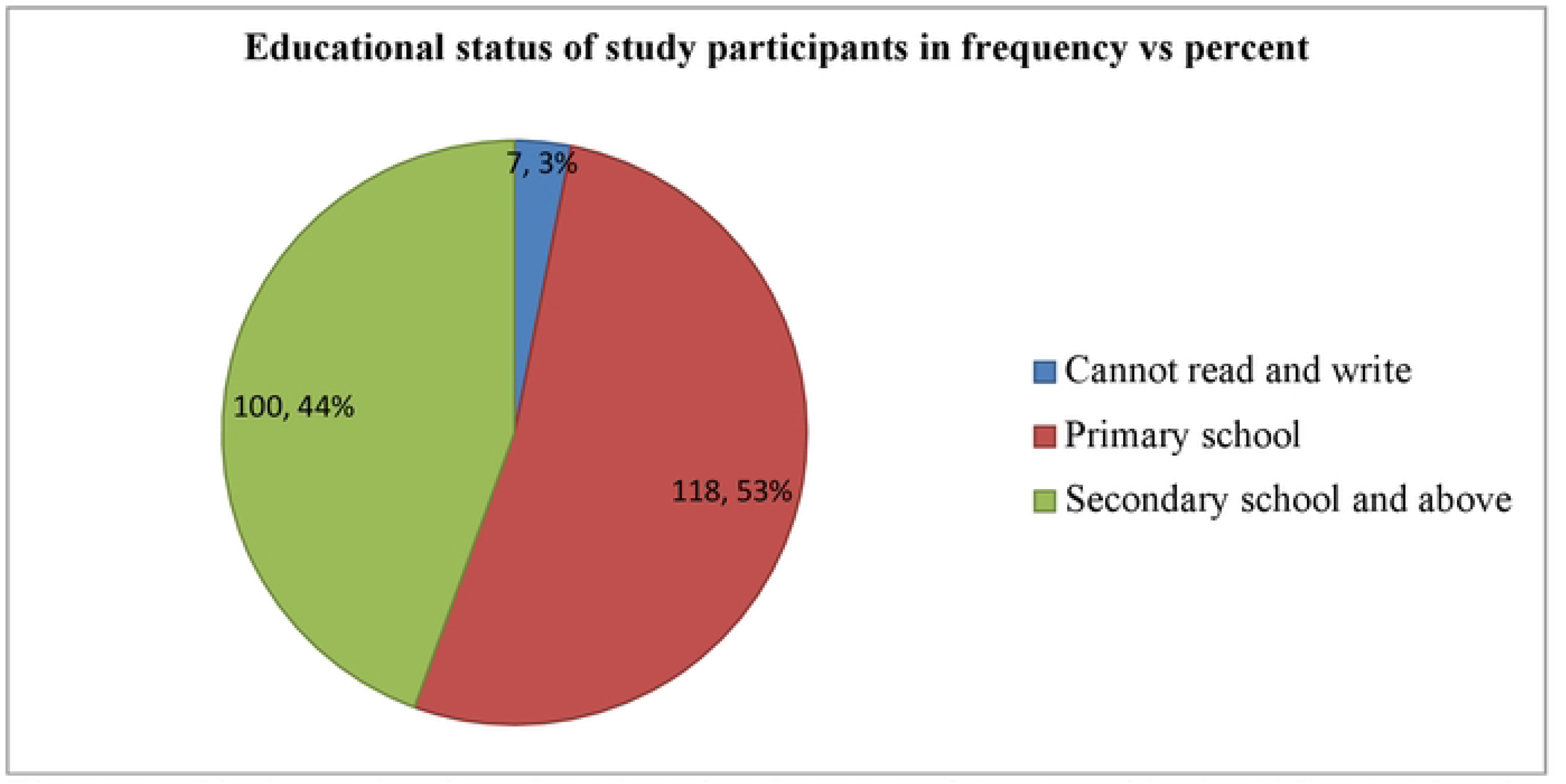
Pi chart showing the educational status of housemaids (n=225) working in communal residences in Jimma City, Southwest Ethiopia, 2022.

### 3.2. Bacterial isolates

The majority, 162(72%) of study participants tested positive for one or more than one bacterial hand contaminant. The total number of isolated bacteria from hand swab samples was 224. *Staphylococcus aureus* 71(31.6%) was the predominant bacterial species isolated from hand swabs of housemaids followed by *Klebsiella species* 52(23.1%) and *Escherichia coli* 48(21.3%) whereas the least isolated bacteria was *Coagulase-Negative Staphylococci (*0.90%). However, bacteria weren’t isolated from the hand swabs of 63(28%) of the study participants (**Table 2**).

**Table 1:** Zone diameter interpretive standards for the determination of antimicrobial agent sensitivity and resistance test by Disk Diffusion method.

| Antimicrobial agent | Disk content (µg) | Interpretive categories and zone diameter breakpoints nearest whole mm |  |  |
| --- | --- | --- | --- | --- |
|  |  | Sensitive | Intermediate | Resistant |
| Tetracycline | 30 | ≥15 | 12-14 | ≤11 |
| Ceftriaxone | 30 | ≥23 | 20-22 | ≤19 |
| Chloramphenicol | 30 | ≥18 | 13-17 | ≤12 |
| Gentamicin | 10 | ≥15 | 13-14 | ≤12 |
| Vancomycin | 30 | ≥12 | 10-11 | ≤9 |
| Ceftazidime | 30 | ≥21 | 18-20 | ≤17 |

Disk Diffusion method [30].

**Table 2:**
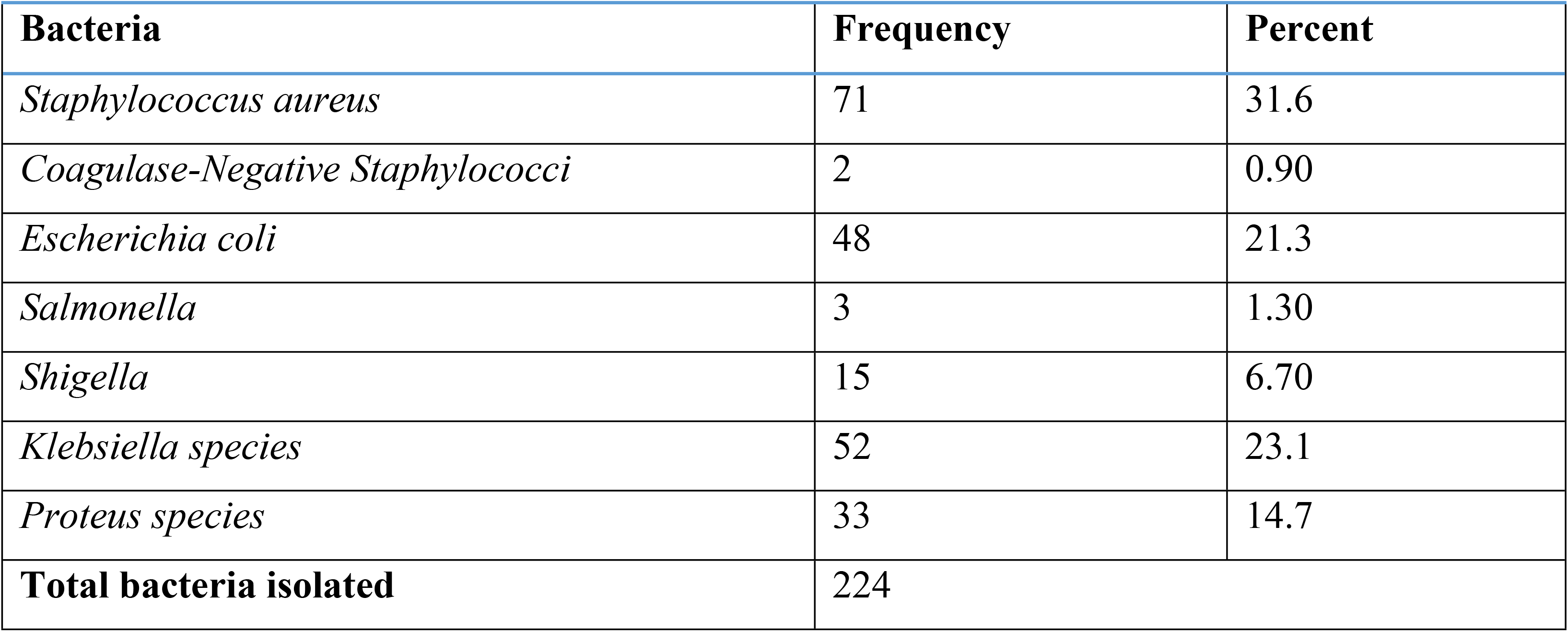
Types of bacteria isolated from hand swabs of housemaids (n=225) working in communal living residences in Jimma City, Southwest Ethiopia, 2022.

| Bacteria | Frequency | Percent |
| --- | --- | --- |
| *Staphylococcus aureus* | 71 | 31.6 |
| *Coagulase-Negative Staphylococci* | 2 | 0.90 |
| *Escherichia coli* | 48 | 21.3 |
| *Salmonella* | 3 | 1.30 |
| *Shigella* | 15 | 6.70 |
| *Klebsiella species* | 52 | 23.1 |
| *Proteus species* | 33 | 14.7 |
| **Total bacteria isolated** | 224 |  |

#### 3.2.1. Risk factors of bacteria

In the present study, different relevant factors were assessed for possible association with bacterial isolation rate from hand swabs among the respondents (**Table 3**).

**Table 3:** Risks factors related to the presence of bacterial isolation from hand swabs of housemaids (n=225) engaged in communal living residences in Jimma City, Southwest Ethiopia, 2022.

| Variables | Category | The bacterial culture results from hand swabs |  | Association |
| --- | --- | --- | --- | --- |
|  |  | Negative n (%) | Positive n (%) |  |
| Age | 18-24 years | 49(26.9) | 133(73.1) | $\chi^2_{(df=2)}= 3.534$<br>$P=.171$ |
|  | 25-30 Years | 9(26.5) | 25(73.5) |  |
|  | ≥31 Years | 5(55.6) | 4(44.4) |  |
| Educational status | Cannot read & write | 1(14.3) | 6(85.7) | $\chi^2_{(df=2)}=1.835$<br>$P=.399$ |
|  | Primary school | 30(25.4) | 88(74.6) |  |
|  | Secondary & above | 32(32.0) | 68(68) |  |
| Fingernail status | Not trimmed | 10(20.4) | 39(79.6) | $\chi^2_{(df=1)}= 1.791$<br>$P=.181$ |
|  | Trimmed | 53(30.1) | 123(69.9) |  |
| Wash hands frequently | No | 2(11.1) | 16(88.9) | $\chi^2_{(df=1)}= 2.768$<br>$P=.096$ |
|  | Yes | 61(29.5) | 146(70.5) |  |
| How to wash your hands? | Only with water | 8(26.7) | 22(73.3) | $\chi^2_{(df=1)}= .133$<br>$P=.716$ |
|  | Water with soap | 53(29.9) | 124(70.1) |  |
| Wash hands frequently with soap/other detergents | No | 3(60.0) | 2(40.0) | $\chi^2_{(df=1)}= 2.216$<br>$P=.137$ |
|  | Yes | 50(29.1) | 122(70.9) |  |
| Follow five steps to wash hands the right way | No | 36(26.5) | 100(73.5) | $\chi^2_{(df=1)}= 1.715$<br>$P=.190$ |
|  | Yes | 25(35.2) | 46(64.8) |  |
| Remove the watch, ring, and bracelet during hand washing | No | 36(24.7) | 110(75.3) | $\chi^2_{(df=1)}= 5.517$<br>$P=.019$ |
|  | Yes | 25(41.0) | 36(59.0) |  |
| Wash hands for 20 seconds | No | 25(23.8) | 80(76.2) | $\chi^2_{(df=1)}= 3.283$<br>$P=.070$ |
|  | Yes | 36(35.3) | 66(64.7) |  |

The number of positive bacterial cultures from hand swabs was higher among the age group of 25–30 years (73.5%) and 18–24 years (73.1%) than their counterparts (*P*=.171). Similarly, the frequency of isolation rate of bacteria from hand swabs of housemaids was higher among those who cannot read and write (85.7%) than those housemaids who attended primary school (74.6%) and secondary school and above (68%) (P=.399) (**Table 3**).

The frequency of isolation rate of bacteria from hand swabs of housemaids was higher among housemaids who didn’t wash their hands frequently (88.9%) than their counterparts (P=.096). In addition, the frequency of the isolation rate of bacteria from hand swabs was higher among housemaids who wash hands frequently with soap/other detergents (70.9%) than those housemaids who didn’t wash their hands frequently with soap/other detergents (40.0%) (P=.137). Furthermore, the isolation rate of bacteria from swabs was higher among housemaids who didn’t follow five steps to wash their hands the right way (73.5%) (P=.190) and remove watch, ring, and bracelet during hand washing (75.3%) (P=.019) than their counterparts (**Table 3**).

The expected risk factors on socio-demographic factors such as age and educational status, and housemaids’ hand hygiene practices such as fingernail status, frequent handwashing, how to wash hands, use of soap and water for frequent hand washing, following five steps to wash hands in the right way and time in second to wash hands weren’t found to be significantly associated with a bacteria isolation rate of hand swabs. However, removing a watch, ring, and bracelet during hand washing was found to be statistically associated (P= P=.019) (**Table 3**).

### 3.3. Antimicrobial susceptibility pattern of bacterial isolates

The majority, 70(98.6%) of *Staphylococcus aureus* were sensitive to Chloramphenicol followed by 51(71.8%) and 46(64.8%) sensitive to Vancomycin and Gentamycin respectively. *Coagulase-Negative Staphylococci* were sensitive to all drugs except a single isolate of *Coagulase-Negative Staphylococci* resistant to Ceftriaxone (**Table 4**).

**Table 4:** Antimicrobial susceptibility pattern of bacteria isolated from hand swabs of housemaids working in communal living residences in Jimma City, Southwest Ethiopia, 2022.

| Bacterial isolate | Total | SP | TE[n(%)] | CRO [n(%)] | C[n(%)] | GEN[n(%)] | VA[n(%)] | CAZ[n(%)] |
| --- | --- | --- | --- | --- | --- | --- | --- | --- |
| *S.aureus* | 71 | S | 42[59.2] | 45[63.4] | 70[98.6] | 46[64.8] | 51[71.8] | 30[42.3] |
|  |  | I | 9[12.7] | 4[5.6] | 1[1.4] | 25[35.2] | 1[1.4] | 25[35.2] |
|  |  | R | 20[28.2] | 22[31.0] | 0[0.00] | 0[0.00] | 19[26.8] | 16[22.5] |
| *CNS* | 2 | S | 2 | 0 | 2 | 2 | 2 | 2 |
|  |  | I | 0 | 1 | 0 | 0 | 0 | 0 |
|  |  | R | 0 | 1 | 0 | 0 | 0 | 0 |
| *E.coli* | 48 | S | 36[75.0] | 38[79.2] | 42[87.5] | 30[62.5] | 36[75.0] | 37[77.1] |
|  |  | I | 0[0.00] | 4[8.30] | 0[0.00] | 18[37.5] | 0[0.00] | 5[10.4] |
|  |  | R | 12[25.0] | 6[12.5] | 6[12.5] | 0[0.00] | 12[25.0] | 6[12.5] |
| *Salmonella* | 3 | S | 1 | 2 | 3 | 1 | 1 | 2 |
|  |  | I | 1 | 0 | 0 | 1 | 0 | 1 |
|  |  | R | 1 | 1 | 0 | 1 | 2 | 0 |
| *Shigella* | 15 | S | 3[20.0] | 8[53.3] | 12[80.0] | 12[80.0] | 0[0.00] | 8[53.3] |
|  |  | I | 5[33.3] | 3[20.0] | 3[20.0] | 0[0.00] | 0[0.00] | 3[20.0] |
|  |  | R | 7[46.7] | 4[26.7] | 0[0.00] | 3[20.0] | 15[100.0] | 4[26.7] |
| *Klebsiella species* | 52 | S | 25[48.1] | 28[53.8] | 46[88.5] | 31[59.6] | 5[9.6] | 28[53.8] |
|  |  | I | 15[28.8] | 14[26.9] | 6[11.5] | 21[40.4] | 23[44.2] | 7[13.5] |
|  |  | R | 12[23.1] | 10[19.2] | 0[0.00] | 0[0.00] | 24[46.2] | 17[32.7] |
| *Proteus species* | 33 | S | 23[69.7] | 9[27.3] | 33[100.0] | 27[81.8] | 16[48.5] | 5[15.2] |
|  |  | I | 1[3.0] | 8[24.2] | 0[0.00] | 5[15.2] | 7[21.2] | 20[60.6] |
|  |  | R | 9[27.3] | 16[48.5] | 0[0.00] | 1[3.0] | 10[30.3] | 8[24.2] |

**CNS:** Coagulase-Negative Staphylococci, **SP:** Sensitivity pattern; **S:** Sensitivity; **I:** Intermediate; **R:** Resistant; **TE:** Tetracycline; **CRO:** Ceftriaxone; **C:** Chloramphenicol; **GEN:** Gentamycin; **VA:** Vancomycin; **CAZ:** Ceftazidime; **n:** Number

More than ⅔, 36(75%), 38(79.2%), 42(87.5%), and 37(77.1%) of *Escherichia coli* were sensitive to Tetracycline, Vancomycin, Ceftriaxone, Chloramphenicol, and Ceftazidime respectively, while no resistance was recorded on Gentamycin with only 18(37.5%) of *Escherichia coli*, were intermediates (**Table 4**).

Thirty-three (100%) and 46(88.5%) of *Proteus species* and *Klebsiella species* were sensitive to Chloramphenicol whereas 16(48.5%) and 24(46.2%) of *Proteus species and Klebsiella species* were resistant to Ceftriaxone and Vancomycin respectively. Despite three *Salmonella* and 12(80.0%) of *Shigella* being sensitive to Chloramphenicol, two *Salmonella* and 15(100%) of *Shigella* were resistant to Vancomycin respectively (**Table 4**).

Antimicrobial susceptibility patterns of different bacteria isolated from swab samples of housemaids working in communal living residences in Jimma City are presented in (**Table 4**).

## 4. Discussion

Housemaids with poor hand hygiene could be potential sources of infection due to pathogenic bacteria which can cause food contamination, and consequently food-borne diseases that pose a potential risk to public health [**22**,**31**]. Due to the scarcity of published information, bacterial contamination level among housemaids in Ethiopia is underexplored. Therefore, the present study was undertaken to assess the profiles of bacteria isolates and associated factors as well as their antimicrobial resistance pattern among housemaids working in communal living residences in Jimma City, Ethiopia.

Among 225 respondents who participated in this study, one or more bacterial pathogens were isolated from 162 study subjects with a prevalence of 72% (95%CI: 66.2 – 77.8%). This result is nearly coherent with the results reported across the globe: Sari city, north of Iran (62.2%) [**31**], Tripoli, Libya (71.41%) [**32**], Alexandria, Egypt (60%) [**33**], Mauritius (91.0%) [**12**] and Ethiopia (49.6%,70.1%,55.7% & 83.9%) [**8–10**,**34**]. On the other hand, the result is higher than the results reported in Eastern India (37.9%) [**5**], Sudan (23.2%) [**7**], and Ethiopia (29.5%) [**14**]. The isolation of bacteria from hand swabs could be due to improper handwashing practices such as not removing watches, rings, and bracelets during handwashing. Evidence from a study indicated that jewelry like a watch, rings, and bracelets would lead to bacterial colonization underneath unless removed and rinsed thoroughly during handwashing [**35**]. In addition, isolation of bacteria illustrates the concept of fecal contamination due to poor hand hygiene [**36**]. On other the hand reason would be likely because of handwashing water quality. Evidences revealed that the bacterial contamination of hands significantly affected by handwashing water [**37**,**38**].

The following bacteria pathogens were isolated *Staphylococcus aureus, Coagulase-Negative Staphylococci, Escherichia coli, Salmonella, Shigella, Klebsiella species*, and *Proteus species. Staphylococcus aureus* (31.8%) was the predominant bacterial species isolated from hand swabs of housemaids followed by *Klebsiella species* (23.3%) and *E*.*coli* (21.5%) respectively. This result coincided with the results reported in previous studies [**7**,**8**,**10**,**14**,**31**,**32**,**34**,**39**]. The isolation of *Staphylococcus aureus* could be because it is pathogenic bacteria that are normal flora of the skin and other body parts whereas the isolation of *E*.*coli, Salmonella, Shigella, Klebsiella species*, and *Proteus species* illustrates the concept of fecal contamination due to poor hand hygiene practices. De Alwis and colleagues revealed that contaminated surfaces such as toilets and washrooms could be the sources of contamination of the hands when a person comes into contact [**36**]. In addition, the microbial contamination of hands could be due to the ineffectiveness of handwashing agents [**40**]. Moreover, ignorance of handwashing with soap, touching dirty materials, and long fingernails [**12**,**41**]. Therefore, the detection of bacteria isolated from the hand of housemaids might pose potential outbreaks for the immediate servant or the community as a whole [**9**]. Due to this, it’s a public health problem of major concern [**42**]. The majority of *Staphylococcus aureus* (98.6%) was sensitive to Chloramphenicol followed by 71.8% and 64.8% to Vancomycin and Gentamycin respectively. This result is lower than the result reported at the University of Gondar Referral Hospital, Ethiopia (76.9%, 100%, and 82.1% were sensitive to Chloramphenicol, Vancomycin, and Gentamycin) [**10**].

In this study more than two-thirds, 75%, 79.2%, 87.5%, and 77.1% of *Escherichia coli* were sensitive to Tetracycline, Vancomycin, Ceftriaxone, Chloramphenicol, and Ceftazidime. In addition, nearly 100% and 88.5% of *Proteus species* and *Klebsiella species* were sensitive to Chloramphenicol. This result is higher than the results reported in the University of Gondar Cafeteria, University of Gondar Referral Hospital, and Debre Markos, Ethiopia [**9**,**10**,**14**].

Despite three *Salmonella* and 80.0% of *Shigella* being sensitive to Chloramphenicol, two *Salmonella* and 100% of *Shigella* were resistant to Vancomycin. This result is higher than the result reported in the University of Gondar Cafeteria [**9**]. In addition, single *Salmonella* and 46.7% of *Shigella* were resistant to Tetracycline. This result is lower than the result reported at Debre Markos University catering establishments [**22**].

Even though a high rate of bacteria’s isolate sensitivity to Chloramphenicol in the present study, a high frequency of drugs resistance to Tetracycline, Vancomycin, Gentamycin, and Ceftriaxone was observed for *Staphylococcus aureus, Coagulase-Negative Staphylococci, Escherichia coli, Salmonella, Shigella, Klebsiella species*, and *Proteus species*. Nowadays, antimicrobial resistance is an emerging global challenge that results in the spread of infectious diseases that affect human populations [**21**,**23**]. Those drug-resistant microbes can multiply, carry on and produce harm because of a complex set of causes: biological processes, human behaviors, and other social factors [**20**]. The resistance to drugs could be because they developed mechanisms (evolutionary processes) or be natural phenomena that microbe tends to adapt it [**19**,**20**]. Another reason could be the inappropriate use of drugs by the community, the use of antibiotics in animals, and the external environment [**21–23**]. In addition, a global connection of a large human population allows microbes into the environment to which all of humanity has access to it [**23**].

In advance, there are many infection control strategies to cope with the results from pathogenic bacteria and their AMR at an individual level as well as at the community level suggested by many scientific communities. Thus, classic communicable disease control methods particularly hand-hygiene remain the cornerstone to curb such public health issues [**43**]. Proper handwashing minimizes the expansion of fecal-oral pathogenic microbes from hands and other sources of the environment [**44**]. Moreover, to prevent and control antimicrobial drug-resistant microbes, washing hands regularly and an improvement in hand hygiene are up to date [**45**]. Therefore, practicing good hand hygiene can reduce outbreaks of pathogen transmission and minimize the spread of antibiotic resistance micro-organisms [**46**].

## 5. Conclusions

This study revealed that 72% of housemaids engaged in dwellings tested positive for one or more than one bacterial contaminant of the hands. The following bacterial pathogens were identified from hand swabs of study subjects: *Staphylococcus aureus, Coagulase-Negative Staphylococci, Escherichia coli, Salmonella, Shigella, Klebsiella species*, and *Proteus species*. Thus, the hands of housemaids could be very important potential sources of disease-causing bacterial pathogens that would result in the potential risk of foodborne diseases. The presence of bacterial pathogens from hand swabs was significantly associated with removing a watch, ring, and bracelet during hand washing. Moreover, most of the isolated bacteria were sensitive to Chloramphenicol while the majority of them were resistant to Tetracycline, Gentamycin, Vancomycin, and Ceftazidime. In advance, there are many infection control strategies to cope with the results from such pathogenic bacteria at an individual level as well as at the community level. In resource-limited settings, regular handwashing and improvement in the hand-hygiene are up to date to prevent and control antimicrobial-resistant microbes. Therefore, housemaids should practice proper hand hygiene to reduce/remove pathogenic bacteria, and minimize the spread of antibiotic-resistant micro-organisms. In addition, any community health worker, regional, national, and stakeholders who are engaged in community health should create awareness about regular handwashing and its relevance in the prevention and reduction of pathogenic micro-organisms from hands in the wider community.

## Data Availability

All relevant data are within the manuscript and its Supporting Information files.

## Acknowledgment

We would like to thank the study participants for being part of the study. In addition, we also gratefully acknowledge the households of study participants, Jimma city municipality, and Jimma University community service directives for their collaboration. Our heartfelt thanks go to data collectors Mr. Dawit Abera, Mr. Bizuwerk Sharew, and Mr. Soressa Gershe. Finally, we are joyful to thank Jimma University, Institute of Health for financial support for the completion of the study.

## Lists of Abbreviations

AMR: Antimicrobial Resistance
EMB: Eosin Methylene Blue
IRB: Institutional Review Board
KIA: Kliger Iron Agar
MSA: Mannitol Salt Agar
NCCLS: National Committee for Clinical Laboratory Standards
SCA: Simon Citrate Agar
SIM: Sulfide Indole Motility
SOP: Standard Operating Procedure
SPSS: Statistical Package for Social Science
SSA: Salmonella-Shigella Agar
PPE: Personal protective equipment

## Author contributions

**Conceptualization**: Tadele Shiwito Ango

**Data curation**: Tadele Shiwito Ango

**Formal analysis**: Tadele Shiwito Ango

**Investigation**: Tadele Shiwito Ango

**Methodology**: Tadele Shiwito Ango

**Software**: Tadele Shiwito Ango

**Visualization**: Tadele Shiwito Ango, Tizita Teshome, Tesfalem Getahun, Girma Mamo, Negalign Biyadgie

**Writing-original draft**: Tadele Shiwito Ango, Girma Mamo, Negalgn Byadgie

**Writing, reviewing & editing**: Tadele Shiwito Ango, Tizita Teshome, Tesfalem Getahun, Girma Mamo, Negalgn Byadgie

